# Effects of home confinement on mental health and lifestyle behaviours during the COVID-19 outbreak: Insight from the “ECLB-COVID19” multi countries survey

**DOI:** 10.1101/2020.05.04.20091017

**Authors:** Achraf Ammar, Khaled Trabelsi, Michael Brach, Hamdi Chtourou, Omar Boukhris, Liwa Masmoudi, Bassem Bouaziz, Ellen Bentlage, Daniella How, Mona Ahmed, Patrick Mueller, Notger Mueller, Asma Aloui, Omar Hammouda, Laisa Liane Paineiras-Domingos, Annemarie Braakman-jansen, Christian Wrede, Sophia Bastoni, Carlos Soares Pernambuco, Leonardo Mataruna, Morteza Taheri, Khadijeh Irandoust, Aïmen Khacharem, Nicola L Bragazzi, Karim Chamari, Jordan M Glenn, Nicholas T Bott, Faiez Gargouri, Lotfi Chaari, Hadj Batatia, Gamal Mohamed Ali, Osama Abdelkarim, Mohamed Jarraya, Kais El Abed, Nizar Souissi, Lisette Van Gemert-Pijnen, Bryan L Riemann, Laurel Riemann, Wassim Moalla, Jonathan Gómez-Raja, Monique Epstein, Robbert Sanderman, Sebastian Schulz, Achim Jerg, Ramzi Al-Horani, Taysir Mansi, Mohamed Jmail, Fernando Barbosa, Fernando Santos, Boštjan Šimunič, Rado Pišot, Donald Cowan, Andrea Gaggioli, Stephen J Bailey, Jürgen Steinacker, Tarak Driss, Anita Hoekelmann

## Abstract

**Background:** Although recognised as effective measures to curb the spread of the COVID-19 outbreak, social distancing and self-isolation, have been suggested to generate burden throughout the population. To provide scientific data to help identify risk-factors for the psychosocial strain during the COVID-19 outbreak, an international cross-disciplinary online survey was circulated in April 2020. This report outlines the mental, emotional and behavioural consequences of COVID-19 home confinement.

**Method:** Thirty-five research organisations from four continents promoted the survey through their networks to the general society, in Ten different languages. Questions were presented in a differential format with questions related to responses “before” and “during” confinement period.

**Results:** 1047 replies (54% women) from Western-Asia (36%), North-Africa (40%), Europe (21%) and other countries (3%) were analysed. The COVID-19 home confinement evoked a negative effect on mental wellbeing and emotional status (P < 0.001; 0.43 ≤ d ≤ 0.65) with a greater proportion of individuals experiencing psychosocial and emotional disorders (10% to 16.5%). These psychosocial tolls were associated with unhealthy lifestyle behaviours with a greater proportion of individuals experiencing (i) physical (+15.2%) and social (71.2%) inactivity, (ii) poor sleep quality (12.8%), (iii) unhealthy diet behaviours (10%), and (iv) unemployment (6%). Conversely, participants demonstrated a greater use (15%) of technology solutions during the confinement period.

**Conclusion:** These findings elucidate the risk of psychosocial strain during the current home confinement period and provide a clear remit for the urgent implementation of technology-based intervention to foster an Active and Healthy Confinement Lifestyle (AHCL).

## Introduction

Coronavirus disease 2019 (COVID-19) is an emerging infectious disease caused by newly discovered Severe Acute Respiratory Syndrome Coronavirus 2 (SARS-CoV-2).^1^ The disease was first identified in December 2019 in Wuhan, the capital of China’s Hubei province and has since spread globally to affect around 3 million people (4^th^ week of April 2020), including nearly 200,000 deaths in more than 220 countries.^2^ Due to the ever-growing number of confirmed cases and to avoid overwhelming health systems, WHO and public health authorities around the world are acting to contain the rapid spread of the COVID-19 outbreak, with primary measures focusing on social distancing, self-isolation, and nationwide lockdowns.

Although recognized with hygiene care as one of the most effective measures to curb the spread of disease, the weakening of social contact result in the devastating loss of leisure and working hours, disruption of normal lifestyle, and generation of stress throughout the population (WHO, 2020b, Hossain, 2020).^3,4^ As a result, anxiety, frustration, panic attacks, loss or sudden increase of appetite, insomnia, depression, mood-swings, delusions, fear, sleep disorders, and suicidal/ domestic-violence cases have become quite common during lockdowns with helpline numbers being overloaded through surges in SOS.^5-8^ Similarly, Brooks et al.^9^ reported several psychological issues during quarantine periods in patients including: emotional and mood disturbance, numbness, depression, irritability, stress, anger, nervousness, guilt, sadness, fear, vigilant handwashing and avoidance of crowd (SARS, H1N1 influenza, Equine influenza and Ebola). During these periods of and precautionary isolation, Purssell et al.^10^ and Sharma et al.^11^ reported negative psychological effects (i.e., increased levels of anxiety and depression). Social impacts have also been reported, including engendered limited visiting, lesser interaction with providers, and social exclusion.^12^

Therefore, in such times of crisis, there exists an emergent need to support mental and psychosocial well-being in target groups during outbreaks to minimize the psychosocial toll. In this context, mental health initiatives focused on (*i*) educating public and health care workers on how to properly deal with the immense pressure and anxiety, (*ii*) providing targeted mental health surveillance followed by effective interventions for at-risk populations (*e.g*., patients with prior mental health diagnosis, the elderly, people in total home confinement), and (*iii*) proactively establishing mental health programs specifically designed to manage the pandemic’s aftermath, have been recently suggested as urgent measures of preventive and early intervention (WHO, 2020b; Galea et al. 2020, Usher et al. 2020).^3,13,14^ The psychosocial needs of at-risk individuals, including those in quarantine and/or home confinement, are suggested to be unique.^14^ Preventive, early and rehabilitation-focused interventions to promote mental wellbeing should be designed to be “crisis-oriented” and should be informed by outcomes from scientific research, as opposed to hypothetical and speculative suggestions. Consistent with this standpoint, a recent “paper advises” article highlighted the urgent need of research to help improve understanding of the mental health consequences of the COVID-19 pandemic on the public.^15^ Therefore, to provide scientific data to help characterise the psychosocial effects of the COVID-19 crisis, our ECLB-COVID19 research group recently launched a multiple-language and multi-country anonymous survey to assess the effects of home confinement on psychosocial health status and multiple lifestyle behaviours during the COVID-19 outbreak (ECLB-COVID19).

An accurate understanding of behavioural changes accompanying the COVID-19 lockdowns is a necessary step toward a crisis-oriented based-research intervention to foster healthy lifestyle and physical and mental wellbeing. Based on data extracted from the first thousand multi-country responses (1047 participants), the present manuscript aims to provide insight into the effect of home-confinement on mental wellbeing, depression, life satisfaction and multidimension lifestyle behaviours (i.e., social participation, physical activity, dietary behaviours, sleep quality and technology-use). Additionally, we aimed at identifying possible relationships between psychosocial and behavioural changes during the confinement period.

We hypothesize that social distancing would negatively affect mental and emotional wellbeing via increases in sedentary activity, social exclusion, decreasing sleep quality and decreased adherence to healthy diet.

There is a common method description in all ECLB-COVID19 papers.

## Methods

We report findings on the first 1047 replies to an international online-survey on mental health and multi-dimension lifestyle behaviours during home confinement (ECLB-COVID19). ECLB-COVID19 was opened on April 1, 2020, tested by the project’s steering group for a period of 1 week, before starting to spread it worldwide on April 6, 2020. Thirty-five research organizations from Europe, North-Africa, Western Asia and the Americas promoted dissemination and administration of the survey. ECLB-COVID19 was administered in English, German, French, Arabic, Spanish, Portuguese, and Slovenian languages (currently provided also in Dutch, Persian and Italian). The survey included sixty-four questions on health, mental wellbeing, mood, life satisfaction and multidimension lifestyle behaviours (physical activity, diet, social participation, sleep, technology-use, need of psychosocial support). All questions were presented in a differential format, to be answered directly in sequence regarding “before” and “during” confinement conditions.

The study was conducted according to the Declaration of Helsinki. The protocol and the consent form were fully approved (identification code: 62/20) by the Otto von Guericke University Ethics Committee.

## Survey development and promotion

The ECLB-COVID19 electronic survey was designed by a steering group of multidisciplinary scientists and academics (i.e., human science, sport science, neuropsychology and computer science) at the University of Magdeburg (principal investigator), the University of Sfax, the University of Münster and the University of Paris-Nanterre, following a structured review of the literature. The survey was then reviewed and edited by Over 50 colleagues and experts worldwide. The survey was uploaded and shared on the Google online survey platform. A link to the electronic survey was distributed worldwide by consortium colleagues via a range of methods: invitation via e-mails, shared in consortium’s faculties official pages, ResearchGate™, LinkedIn™ and other social media platforms such as Facebook™, WhatsApp™ and Twitter™. Public were also involved in the dissemination plans of our research through the promotion of the ECLB-COVID19 survey in their networks. The survey included an introductory page describing the background and the aims of the survey, the consortium, ethics information for participants and the option to choose one of seven available languages (English, German, French, Arabic, Spanish, Portuguese, and Slovenian). The present study focusses on the first thousand responses (i.e., 1047 participants), which were reached on April 11, 2020, approximately one-week after the survey began. This survey was open for all people worldwide aged 18 years or older. People with cognitive decline are excluded.

## Data privacy and consent of participation

During the informed consent process, survey participants were assured all data would be used only for research purposes. Participants’ answers are anonymous and confidential according to Google’s privacy policy (https://policies.google.com/privacy?hl=en). Participants don’t have to mention their names or contact information. In addition, participant can stop participating in the study and can leave the questionnaire at any stage before the submission process and their responses will not be saved. Response will be saved only by clicking on “submit” button. By completing the survey, participants are acknowledging the above approval form and are consenting to voluntarily participate in this anonymous study. Participants have been requested to be honest in their responses.

## Survey questionnaires

The ECLB-COVID19 is a multi-country electronic survey designed to assess change in multiple lifestyle behaviours during the COVID-19 outbreak. Therefore, a collection of validated and/or crisis-oriented briefs questionnaires were included. These questionnaires assess mental wellbeing (Short Warwick-Edinburgh Mental Well-being Scale (SWEMWBS)),^16^ mood and feeling (Short Mood and Feelings Questionnaire (SMFQ)),^17^ life satisfaction (Short Life Satisfaction Questionnaire for Lockdowns (SLSQL), social participation (Short Social Participation Questionnaire for Lockdowns (SSPQL)), physical activity (International Physical Activity Questionnaire Short Form (IPAQ-SF)),^18^ ^19^ diet behaviours (Short Diet Behaviours Questionnaire for Lockdowns (SDBQL)), sleep quality (Pittsburgh Sleep Quality Index (PSQI))^20^ and some key questions assessing the technology-use behaviours (Short Technology-use Behaviours Questionnaire for Lockdowns (STBQL)), demographic information and the need of psychosocial support. Reliability of the shortened and/or newly adopted questionnaires was tested by the project steering group through piloting, prior to survey administration. These brief crisis-oriented questionnaires showed good to excellent test-retest reliability coefficients (r = 0.84-0.96). A multi-language validated version already existed for the majority of these questionnaires and/or questions. However, for questionnaires that did not already exist in multi-language versions, we followed the procedure of translation and backtranslation, with an additional review for all language versions from the international scientists of our consortium. Detailed descriptions of the aforementioned tools including total score calculation and interpretation of each questionnaires are available as supplementary file 1. As a result, a total of 64 items were included in the ECLB-COVID19 online survey in a differential format. Each item or question requested two answers, one regarding the period before and the other regarding the period during confinement. Thus, participants were guided to compare the situations.

Given the large number of included questions and in order to give a multidimension overview of the recorded change “during” compared to “before” the confinement period, the present paper focuses only on the total scores of the included questionnaires, without detailed analysis regarding specific changes in each questionnaire.

## Data Analysis

Descriptive statistics were used to define the proportion of responses for each question and the total distribution of the total score of each questionnaire. All statistical analyses were performed using the commercial statistical software STATISTICA (StatSoft, Paris, France, version 10.0). Normality of the data distribution was confirmed using the Shapiro-Wilks-W-test. Values were computed and reported as mean ± SD (standard deviation). To assess significant difference in total scored responses between “before” and “during” confinement period, Paired samples t-tests were used for normally distributed data and the Wilcoxon test was used when normality was not assumed. Effect size (Cohen’s d) was calculated to determine the magnitude of the change score and interpreted using the following criteria: 0.2 (small), 0.5 (moderate), and 0.8 (large).^21^ Pearson product-moment correlation tests were used to assess possible relationships between the “before-after” ∆ of the assessed multidimension total scores. Statistical significance was identified at p<0.05.

## Results

### Sample description

1047 participants were included in the preliminary sample. Overall, 54% of the sample were women and 46% were men. Geographical breakdowns were from Asian (36%, mostly from Western Asia), African (40%, mostly from North Africa), European (21%) and other (3%) countries. Age, health status, employment status, level of education, and marital status are presented in Table 1.

**Table 1.** Demographic characteristics of the participants (N = 1047)

| **Variables** |  | **N** | **(%)** |
| --- | --- | --- | --- |
| **Gender** |  |  |  |
|  | Male | 484 | (46.2%) |
|  | Female | 563 | (53.8%) |
| **Continent** |  |  |  |
|  | North Africa | 419 | (40%) |
|  | Western Asia | 377 | (36%) |
|  | Europe | 220 | (21%) |
|  | Other | 31 | (3%) |
| **Age** |  |  |  |
|  | 18-35 | 577 | (55.1%) |
|  | 36-55 | 367 | (35.1%) |
|  | >55 | 103 | (9.8%) |
| **Level of Education** |  |  |  |
|  | Master/doctorate degree | 527 | (50.3%) |
|  | Bachelor's degree | 397 | (37.9%) |
|  | Professional degree | 28 | (2.7%) |
|  | High school graduate, diploma or the equivalent | 69 | (6.6%) |
|  | No schooling completed | 26 | (2.5%) |
| **Marital status** |  |  |  |
|  | Single | 455 | (43.5%) |
|  | Married/Living as couple | 562 | (53.7%) |
|  | Widowed/Divorced/Separated | 30 | (2.9%) |
| **Employment status** |  |  |  |
|  | Employed for wages | 538 | (51.4%) |
|  | Self-employed | 74 | (7.1%) |
|  | Out of work/Unemployed | 75 | (7.2%) |
|  | A student | 259 | (24.7%) |
|  | Retired | 23 | (2.2%) |
|  | Unable to work | 9 | (0.9%) |
|  | Problem caused by COVID-19 | 59 | (5.6%) |
|  | Other | 10 | (1%) |
| **Health state** |  |  |  |
|  | Healthy | 956 | (91.3%) |
|  | With risk factors for cardiovascular disease | 81 | (7.7%) |
|  | With cardiovascular disease | 10 | (1%) |

### Mental wellbeing, depression, life satisfaction and need of psychosocial support

Change in the total score of the of the SWEMWBS, SMFQ, and SLSQL questionnaire and the psychological support key question from “before” to “during” home confinement period are presented in Figure 1. Statistical analysis showed a significant difference in all tested parameters (14.12≤ t ≤ 21.05; P < 0.001, 0.43 ≤ d ≤ 0.65). Particularly, total score in mental wellbeing and life satisfaction questionnaires decreased by 9.4% (t=18.82, p<0.001, d=0.58) and 16% (t=21.05, p<0.001, d=0.65), respectively from “before” to “during” with more individuals (+12.89%) reporting a very low-low mental wellbeing and more people feeling dissatisfied (extremely-slightly) (+16.5%) “during” compared to “before” the confinement period. In contrast, total score in the depression monitoring questionnaire, as well as in the need of psychosocial support question, increased by 44.9% (t=14.12, p<0.001, d=0.43) and 20.2 % (t=14.83, p<0.001, d=0.56) from “before” to “during,” with more people developing depressive symptoms/states (10%) and more people declaring a need (sometimes-all rimes) of psychosocial support (16.1%) “during” compared to “before” the confinement period.

**Figure 1.**
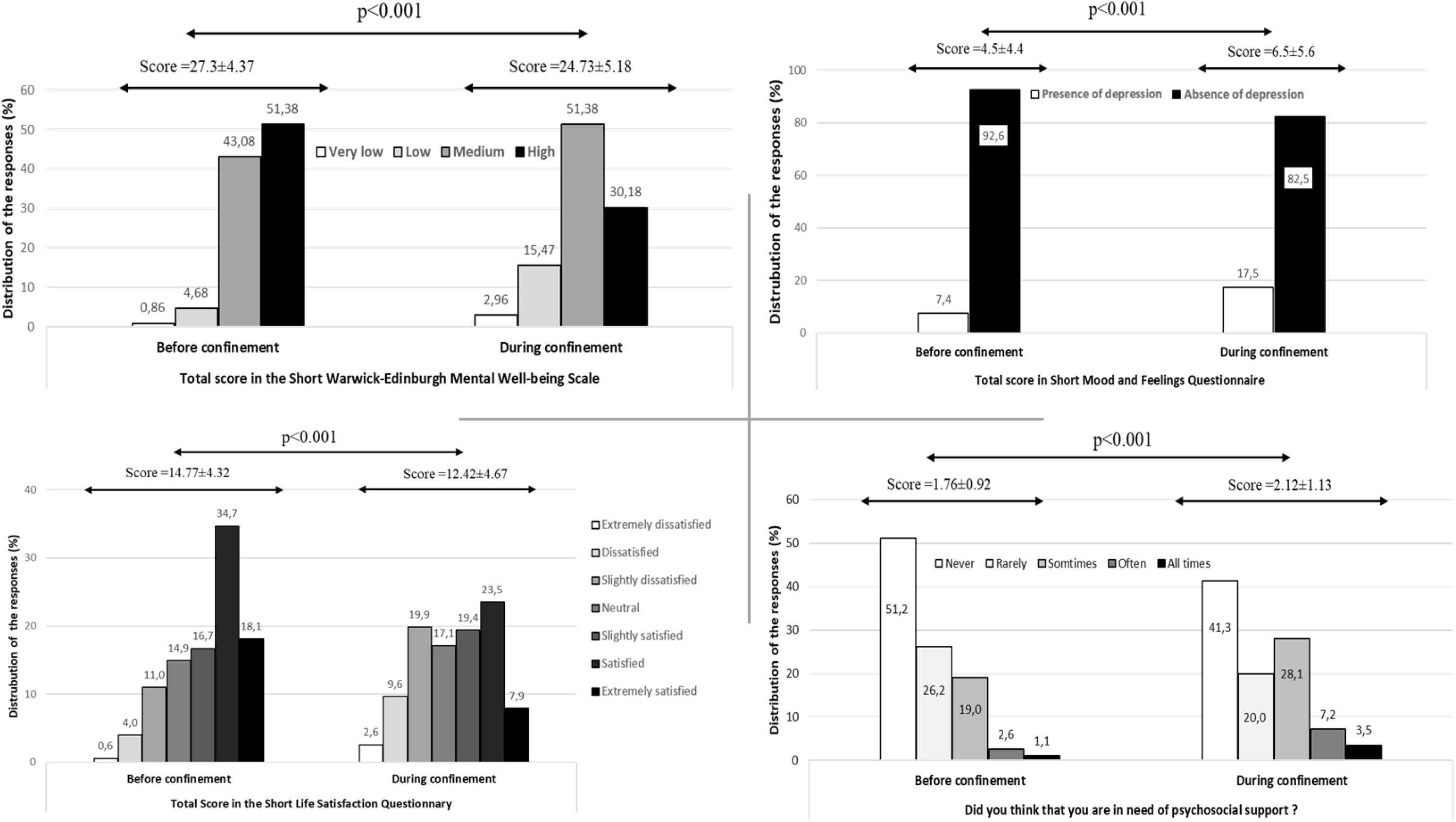
Response to the psychological support key question and total score of the mental wellbeing, mood and feelings, and short life satisfaction questionnaires before and during home confinement.

### Social participation, physical activity, diet and sleep behaviours

Change in the total score of the of the SSPQL, IPAQ-SF, SDBQL, and PSQI questionnaires from “before” to “during” home confinement period are presented in Figure 2. Statistical analysis showed a significant difference between both periods in all tested parameters (10.66≤ t ≤ 69.16; P < 0.001, 0.3 ≤ d ≤ 2.14). Total score in social participation and physical activity (i.e., days/week of all physical activity) questionnaires decreased by 42% (t=69.19 p<0.001, d=2.14) and 24% (t= 15.61, p<0.001, d=0.482), respectively from “before” to “during,” There were more socially (+71.15%, Never-Rarely socially active) and physically (+15.2, 0-1 days/week of all physical activity) inactive individuals “during” compared to “before” the confinement period. In contrast, total score in the diet and sleep monitoring questionnaires increased significantly by 4.4% (t=-10.66, p<0.001, d=0.50) and 12 % (z=10.58, p<0.001, d=0.3) from “before” to “during” with more people experiencing poor sleep quality (+12.8%) and more people classifying (most of the time-always) their diet behaviours as unhealthy (10%) “during” compared to “before” the confinement period.

**Figure 2.**
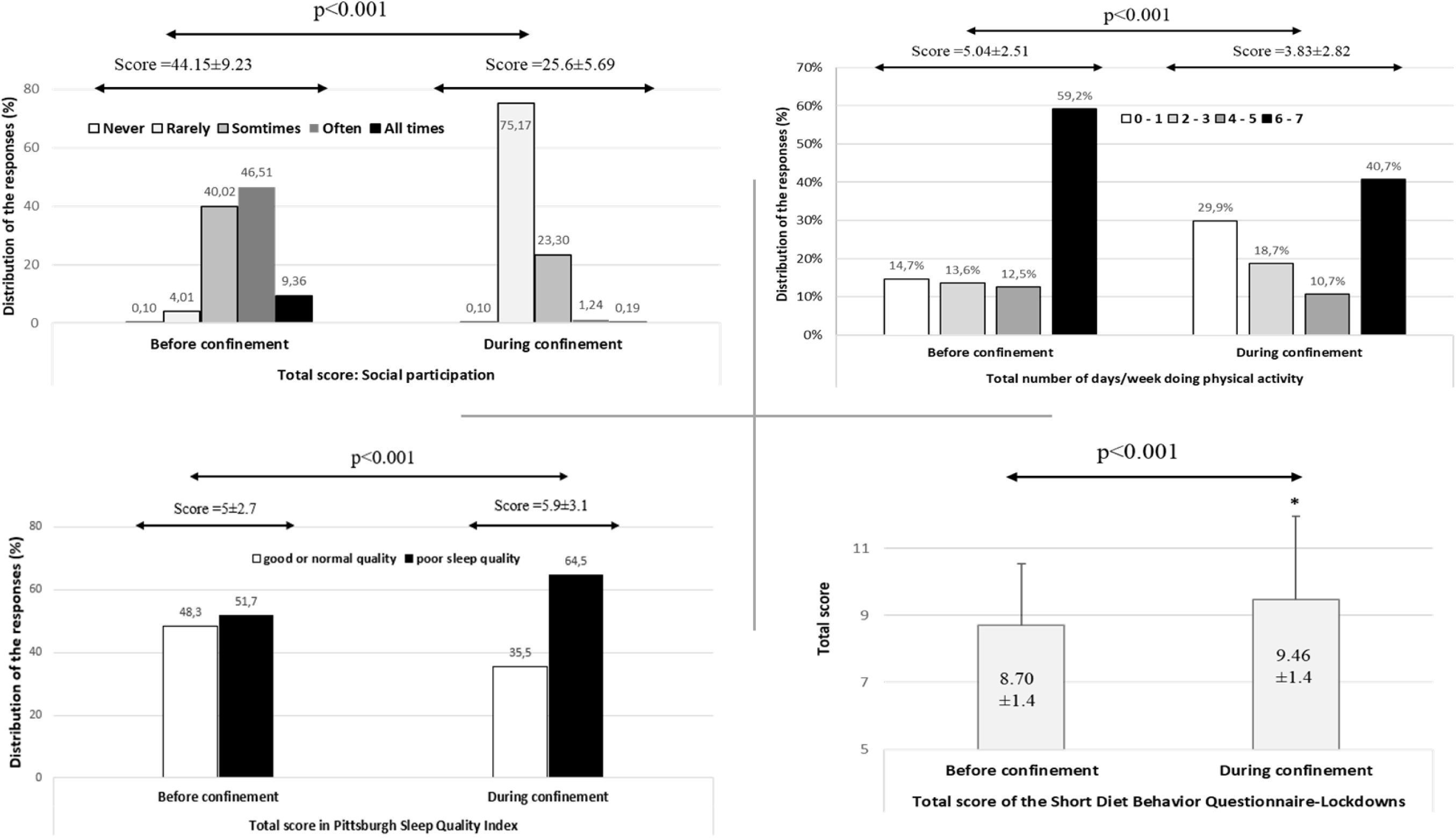
Total score of the social participation, physical activity, diet and sleep behaviors questionnaires before and during home confinement.

### Short Technology-use Lockdowns Questionnaire

Change in technology-use score from “before” to “during” confinement period in response to SLSQL are presented in Figure 3. Statistical analysis showed the total score of the technology-use behaviour increased significantly (8.8%) at “during” compared to “before” home confinement (t= 14.01, P<0.001, d=0.43). Particularly, scores related to the use of internet/social media for communication significantly increased “during” compared to “before” the confinement period t=17.03, P<0.001 and d=0.54. Similarly, higher scores related to the use of technology-based tools for physical activity was registered during the confinement period (t=9.03, p<0.001, d=0.28). However, no significant change was recorded for scores related to the use of technology-based tools for dietary purposes (t=0.61, p=0.53, d=0.01).

**Figure 3.**
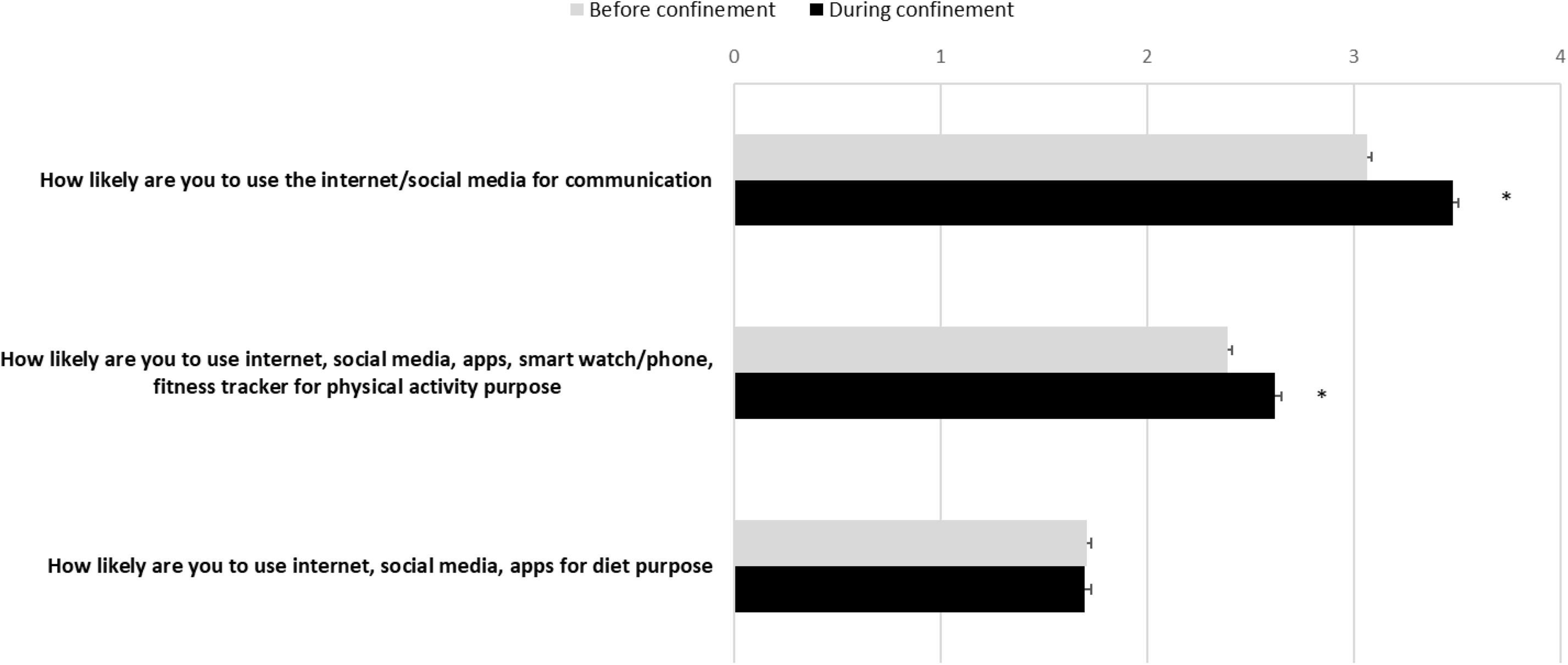
Responses to the Short Technology-use Lockdowns Questionnaire before and during home confinement. Values were computed and reported as mean ± SEM (standard error of the mean). *Significantly different from before confinement at p<0.05.

### Relationship between change in mental and emotional wellbeing and behavioural factors

Table 2 shows the relationship between the change “before-after” of the assessed variables. As this table indicates, the mental and emotional related variables were significantly correlated to the majorities of lifestyle behaviours (0.01 ≤ P ≤ 0.001 and 0.1 ≤ r ≤ 0.41). Particularly, ∆ in total score of mood and feeling questionnaires showed significant correlations to all behavioural changes with positive correlation to the diet and sleep behaviours (p<0.001, 0.3 ≤ r ≤ 0.41) and negative correlation to social participation and physical activity (p<0.001, −0.25 ≤ r≤ −0.14). Inversely, ∆ in total score of mental wellbeing and life satisfaction were positive correlated to social participation (p<0.001, 0.23 ≤ r≤ 0.28) and physical activity (p<0.01, 0.10 ≤ r≤ 0.15) and negatively correlated to the diet (p<0.001, −0.21 ≤ r ≤ −0.14) and sleep behaviours (p<0.001, −0.32 ≤ r ≤ −0.23).

**Table 2.**
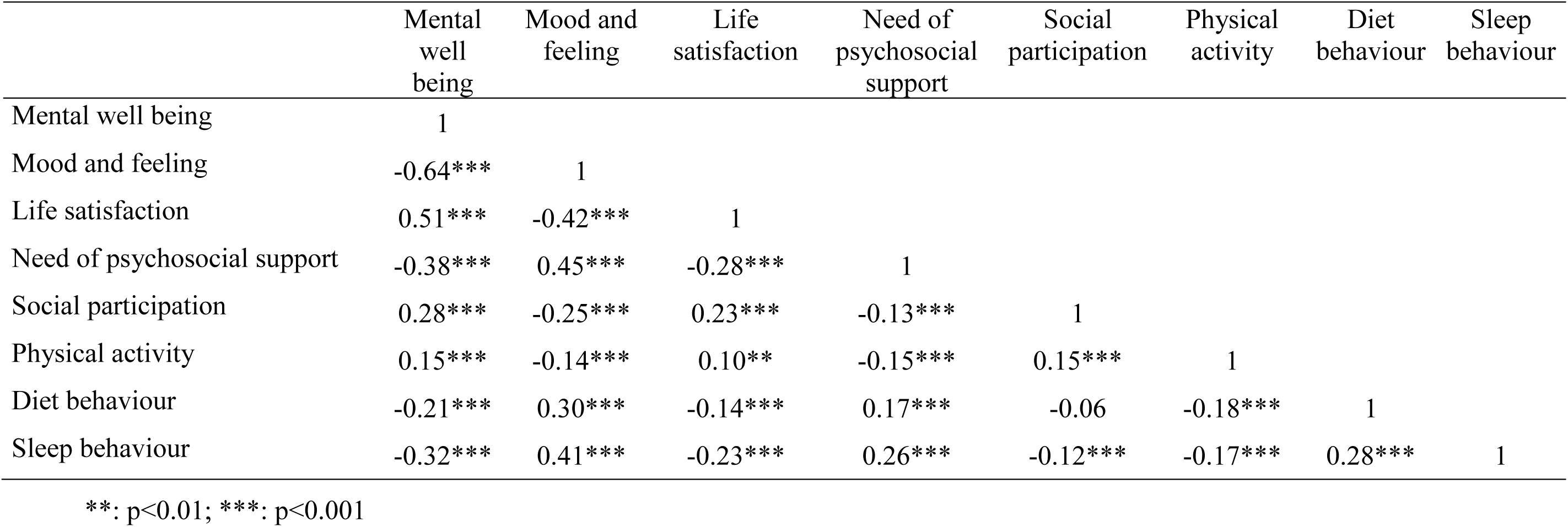
Relationship between delta total score in mental wellbeing, mood and feeling, life satisfaction and the multidimension lifestyle behaviours (social participation, physical activity, diet and sleep)

## Discussion

The present study reports preliminary results from our first 1047 participants (54% female) who responded to our ECLB-COVID19 multiple languages online survey. Preliminary findings from this survey showed COVID-19 home confinement has a negative effect on mental wellbeing and emotional status with more individuals (i) perceiving low mental wellbeing (+12.89%), (ii) feeling dissatisfied (+16.5%), (iii) developing depression (+10%), and (iv) declaring a need of psychosocial support (+16.1%) compared to “before” the confinement period. During similar pandemic crises (2002-2004 SARS outbreak), previous research revealed several negative effects of quarantine measures on mental health and were associated with psychological and emotional problems such as depression and anxiety.^22, 23^ These negative effects (i.e., increased levels of anxiety and depression) have also been reported in two recent systematic reviews and meta-analyses conducted by Purssell et al.^10^ and Sharma et al.^11^ assessing the impact of isolation precaution on quality of life. Similarly, in their recent review of the evidence, Brooks et al.^9^ reported several psychological perturbations and emotional/ mood disturbances such as numbness, depression, irritability, stress, anger, nervousness, guilt, sadness, fear, vigilant handwashing, and avoidance of crowds in infected patients (SARS, MERS, H1N1 influenza, Ebola, and Equine influenza) during quarantine periods. Similarly, results from Chinese studies indicate the COVID-19 outbreak engendered anxiety, depression, sleep problems, and other psychological problems.^7, 8^ This is related to the coupling of psychomental well-being to regular physical activity and to the related effects on immune function.^24^ With significant negative effects of the current COVID-19 pandemic on mental wellbeing, life satisfaction, and depression scores of 1047 participants from different continents, the present findings support these suggestions and elucidate the risk of mental disorders (e.g. low wellbeing, dissatisfaction and depression) during the current home confinement period.

The resultant weakening of social contact with the disruption of normal lifestyles during the COVID-19 outbreaks, have been recently suggested to generate stress throughout the population and thereby to engender lower mental and emotional wellbeing (WHO, 2020b, Gammon and Hunt, 2018).^3, 12^ To provide scientific evidence and deeper the understanding for these suggestions, the present multi-dimension survey also focused on the lifestyle behavioural changes during the COVID-19 outbreak. Main findings showed the negative psycho-emotional effect of COVID-19 home confinement was accompanied with a negative effect on the majority of assessed lifestyle behaviours with more (i) physically inactive peoples (+15.2), (ii) socially isolated peoples (+71.15%), (iii) unemployed peoples (+6%), (iv) more peoples experiencing poor sleep quality (+12.8%), and (v) unhealthy diet behaviours (+10%) compared to “before” the confinement period. Likewise, there are increased number of peoples (+15%) who are “All times” using technology.

These preliminary findings confirm our hypotheses related to the lifestyle behaviours. To better understand the behavioural changes recognized as risk factors of declined psychosocial wellbeing during the confinement period, a correlation analysis between the ∆ change in total scores of all assessed variables from “before” to “during” confinement was performed. Main findings indicate changes in mental wellbeing, mood and feeling and life satisfaction were significantly correlated to changes in lifestyle behaviours, including social participation, physical activity, diet, and sleep. These results suggest low mental wellbeing and life dissatisfaction and high level of depressive symptoms are related to social isolation, sedentary lifestyle, unhealthy diet behaviour and poor sleep quality. Therefore, in order to mitigate the negative physical and psychosocial effects of home confinement, an implementation of a multi-dimension “need-oriented” intervention is warranted. This intervention should focus on enhancing social participation and promoting physical activity (e.g., the German example: allowing people to do outdoor physical activity in the large public garden with respecting distancing and hygiene precautions), healthy food, and sleep quality.

Since participants demonstrated a higher acceptance rate (21.8% vs. 36.8%) toward the use of technology solutions, it seems interesting to foster social communication, and physical and mental wellbeing through technology facilities (e.g., social platform, gamification, *m*health, interactive coach etc.). Indeed, such ICT-based solutions would facilitate the delivery of COVID19-related health services, as well as preventive and rehabilitation crisis-oriented intervention in the communities with a specific challenge to reach risk populations.

To expand the target group, WHO and the national authorities are encouraged to implement, during lockdowns crises, a “Technology-use” support system including factors such as reducing internet fee, providing free based-ICT social inclusion platforms, promoting Gamification, Communication and interactive coaching technologies, tracking contacts and symptoms, switching from 4G to 5G network, to name a few.

### Strengths and limitations

This is the first interdisciplinary international research project evaluating the psychosocial and behavioural changes “during” compared to “before” the home confinement period using a multiple-languages online survey. Preliminary findings from this study offers some important insights into the effect of home confinement on mental wellbeing, emotional health status and the associated multidimension behavioural change in response to the COVID-19 outbreak. However, given that data of the present study has been collected from a heterogeneous population with no criteria-based subsamples analysis, the present findings need to be interpreted with caution. Additionally, since the ECLB-COVID19 survey is still open and meanwhile also available in Dutch, Persian and Italian languages, future post-hoc studies in a more representative sample will be conducted to assess the interaction between the psychosocial strain evoked by COVID-19 and the geographical, demographical, cultural and health characteristics of the participants.

## Conclusion

The preliminary results of the survey reveal a considerable burden for mental wellbeing combined with an unhealthy lifestyle during, compared to before, the confinement enforced by the COVID-19 pandemic. In particular, social and physical inactivity, an unhealthy diet and poor sleep quality were associated with lower mental and emotional wellbeing (i.e., depressive and dissatisfaction feelings) were triggered by the enforced home confinement. These multidimensional negative effects underscore the importance to stakeholders and policy makers to develop, implement and publicise interdisciplinary interventions to mitigate the physical and psychosocial strain evoked by this pandemic. Promoting wellbeing by encouraging individuals to engage in indoor and/or outdoor physical activity in large public parks, whilst conforming with distancing and hygiene recommendations, can be suggested as preliminary measure with evidence for physical and mental benefits. Moreover, since participants have demonstrated a higher acceptance of the use of technology solutions during the confinement period, fostering an Active and Healthy Confinement Lifestyle (AHCL) via an ICT-based approach can be implemented.

A proposed psychosocial strain mitigation strategy from ECLB-COVID19 consortium can be found in the supplementary file 2.

## Data Availability

Data are available from the corresponding author on reasonable request

## Acknowledgement

We thank our consortium’s colleagues who provided insight and expertise that greatly assisted the research. We thank all colleagues and peoples who believed on this initiative and helped to distribute the anonymous survey worldwide. We are also immensely grateful to all participants who #StayHome & #BoostResearch by voluntarily taken the #ECLB-COVID 19 survey.

## References

1. WHO. Naming the coronavirus disease (COVID-19) and the virus that causes it”. World Health Organization. Archived from the original on 28 February 2020. Retrieved 28 February 2020.

2. EDCD. Situation update worldwide, as of 19 April 2020. https://www.ecdc.europa.eu/en/geographical-distribution-2019-ncov-cases. Retrieved 19 April 2020.

3. WHO. Mental health and psychosocial considerations during the COVID-19 outbreak. World Health Organization. https://www.who.int/docs/default-source/coronaviruse/mental-health-considerations.pdf. Retrieved 12 April 2020.

4. Hossain MM, Sultana A, Purohit N. Mental health outcomes of quarantine and isolation for infection prevention: A systematic umbrella review of the global evidence. *Available at SSRN 3561265*. 2020

5. Reuters investigates. COVID’s Other Causalities. https://www.reuters.com/investigates/special-report/health-coronavirus-usa-cost/ Retrieved 12 April 2020.

6. The New York Times. A New Covid-19 Crisis: Domestic Abuse Rises Worldwide. https://www.nytimes.com/2020/04/06/world/coronavirus-domestic-violence.html. Retrieved 12 April 2020.

7. Qiu J, Shen B, Zhao M, Wang Z, Xie B, Xu Y. A nationwide survey of psychological distress among Chinese people in the COVID-19 epidemic: implications and policy recommendations. Gen. Psychiatr 2020; 33, e100213. doi:10.1136/gpsych-2020-100213.

8. Wang C, Pan R, Wan X, Tan Y, Xu L, Ho CS, Ho RC. Immediate Psychological Responses and Associated Factors during the Initial Stage of the 2019 Coronavirus Disease (COVID-19) Epidemic among the General Population in China. Int. J. Environ. Res. Public Health 2020; 17. doi:10.3390/ijerph17051729.

9. Brooks SK, Webster RK, Smith LE, Woodland L, Wessely S, Greenberg N, Rubin GJ. The psychological impact of quarantine and how to reduce it: rapid review of the evidence. The Lancet 2020; 10227: 912–920. https://doi.org/10.1016/S0140-6736(20)30460-8

10. Purssell E, Gould D, Chudleigh J. Impact of isolation on hospitalised patients who are infectious: systematic review with meta-analysis. BMJ open 2020; 10(2). http://dx.doi.org/10.1136/bmjopen-2019-030371

11. Sharma A, Pillai DR, Lu M, Doolan C, Leal J, Kim J, Hollis A. Impact of isolation precautions on quality of life: a meta-analysis. J. Hosp. Infect. Available online 12 February 2020. https://doi.org/10.1016/j.jhin.2020.02.004

12. Gammon J, Hunt J. Source isolation and patient wellbeing in healthcare settings. Br J Nurs 2018; 27(2): 88–91. https://doi.org/10.12968/bjon.2018.27.2.88

13. Galea S, Merchant RM, Lurie N. The Mental Health Consequences of COVID-19 and Physical Distancing: The Need for Prevention and Early Intervention. JAMA Intern Med. 2020. doi:10.1001/jamainternmed.2020.1562.

14. Usher K, Durkin J, Bhullar N. The COVID-19 pandemic and mental health impacts. Int J Ment Health Nurs. 2020. doi:10.1111/inm.12726.

15. Mahase E. Covid-19: Mental health consequences of pandemic need urgent research, paper advises. BMJ 2020; Published 16 April 2020. doi:https://doi.org/10.1136/bmj.m1515

16. Ng Fat L, Scholes S, Boniface S, Mindell J, Stewart-Brown S. Evaluating and establishing the national norms for mental well-being using the short Warwick-Edinburgh Mental Well-being Scale (SWEMWBS): findings from the Health Survey for England. Qual. Life Res. 2017; 26(5): 1129–1144. https://doi.org/10.1007/s11136-016-1454-8

17. Thabrew H, Stasiak K, Bavin LM, Frampton C, Merry S. Validation of the Mood and Feelings Questionnaire (MFQ) and Short Mood and Feelings Questionnaire (SMFQ) in New Zealand help-seeking adolescents. INT J METH PSYCH RES 2018; 27(3): 1–9. https://doi.org/10.1002/mpr.1610

18. Graig CL, Marshall AL, Sjöström M, Bauman AE, Booth ML, Ainsworth BE, et al. International Physical Activity Questionnaire: 12-Contry Reliability and Validity. Med Sci Sports Exerc. 2003; 35(8): 1381–1395.

19. Lee PH, Macfarlane DJ, Lam TH, Stewart SM. Validity of the international physical activity questionnaire short from (PAQ-SF): A systematic review. Int J Behov Nutr Phys Act. 2011; 8: 115.

20. Buysse DJ, Reynolds CF, Monk TH, Berman SR, et al. The Pittsburgh Sleep Quality Index: A new instrument for psychiatric practice and research. Psychiatry Research. 1989; 28: 193–213.

21. Cohen J. The effect size. Statistical power analysis for the behavioral sciences 1988; 77-83.

22. Hawryluck L, Gold WL, Robinson S, Pogorski S, Galea S, Styra, R. SARS control and psychological effects of quarantine, Toronto, Canada. Emerging Infect. Dis. 2004; 10: 1206–1212. doi:10.3201/eid1007.030703.

23. Reynolds DL, Garay JR, Deamond SL, Moran MK, Gold W, Styra R. Understanding, compliance and psychological impact of the SARS quarantine experience. Epidemiol. Infect. 2008; 136: 997–1007. doi:10.1017/S0950268807009156.

24. Alack K, Pilat C, Krüger K. Current knowledge and new challenges in exercise immunology. Dtsch Z Sportmed. 2019; 70: 250–260. Doi: 10.5960/dzsm.2019.391.

25. Collins J, Gibson A, Parkin S, Parkinson R, Shave D, Dyer C. Counselling in the workplace: How time-limited counselling can effect change in well-being. Couns Psychother Res 2012; 12(2): 84–92. https://doi.org/10.1080/14733145.2011.638080.

26. Maheswaran H, Weich S, Powell J, Stewart-Brown S. Evaluating the responsiveness of the Warwick Edinburgh Mental Well-Being Scale (WEMWBS): Group and individual level analysis. Health Qual. Life Outcomes 2012; 10(1): 156. https://doi.org/10.1186/1477-7525-10-156

27. Costello EJ, Angold A. Scales to assess child and adolescent depression: checklists, screens, and nets. J AM ACAD CHILD PSY 1988; 27(6): 726–737. https://doi.org/10.1097/00004583-198811000-00011.

28. Angold A, Costello EJ, Messer SC, Pickles A, Winder F, Silver D. Development of a short questionnaire for use in epidemiological studies of depression in children and adolescents 1995; 5: 237–249.

29. Densley K, Davidson S, Gunn J M. Evaluation of the Social Participation Questionnaire in adult patients with depressive symptoms using Rasch analysis. Qual. Life Res. 2013; 22(8): 1987–1997. https://doi.org/10.1007/s11136-013-0354-4.

